# Disentangling visual exploration differences in cognitive impairment

**DOI:** 10.1101/2023.05.17.23290054

**Authors:** Zifan Jiang, Salman Seyedi, Kayci L. Vickers, Cecelia M. Manzanares, James J. Lah, Allan I. Levey, Gari D. Clifford

## Abstract

**Objective:** Compared to individuals without cognitive impairment (CI), those with CI exhibit differences in both basic oculomotor functions and complex viewing behaviors. However, the characteristics of the differences and how those differences relate to various cognitive functions have not been widely explored. In this work we aimed to quantify those differences and assess general cognitive impairment and specific cognitive functions.

**Methods:** A validated passive viewing memory test with eyetracking was administered to 348 healthy controls and CI individuals. Spatial, temporal, semantic, and other composite features were extracted from the estimated eye-gaze locations on the corresponding pictures displayed during the test. These features were then used to characterize viewing patterns, classify cognitive impairment, and estimate scores in various neuropsychological tests using machine learning.

**Results:** Statistically significant differences in spatial, spatiotemporal, and semantic features were found between healthy controls and individuals with CI. CI group spent more time gazing at the center of the image, looked at more regions of interest (ROI), transitioned less often between ROI yet in a more unpredictable manner, and had different semantic preferences. A combination of these features achieved an area under the receiver-operator curve of 0.78 in differentiating CI individuals from controls. Statistically significant correlations were identified between actual and estimated MoCA scores and other neuropsychological tests.

**Conclusion:** Evaluating visual exploration behaviors provided quantitative and systematic evidence of differences in CI individuals, leading to an improved approach for passive cognitive impairment screening.

**Significance:** The proposed passive, accessible, and scalable approach could help with earlier detection and a better understanding of cognitive impairment.

## I. Introduction

Alzheimer’s disease (AD) is a neurodegenerative disease that typically presents with memory loss and difficulty in in performing cognitive functions, including learning and memory [1]. Mild Cognitive Impairment (MCI) due to AD is a prodromal stage in the AD continuum, when cognitive problems are detected clinically in the absence of any functional decline.

Assessing objective cognitive impairment with neuropsychological measures is an important clinical criterion for diagnosing MCI and dementia [1]. A widely used general cognitive screening tool is the Montreal Cognitive Assessment (MoCA) [2], which briefly assesses several cognitive domains, including executive functioning, immediate and delayed memory, visuospatial abilities, attention, working memory, language, and orientation to time and place. Other neuropsychological tests that are more sensitive to specific cognitive functions have also been widely used, such as the Benson Complex Figure Copy [3] for visual memory and digit span test [4] for attention and working memory. Although effective in assessing cognitive functions, neuropsychological measures can be time-intensive, must be administered, scored, and interpreted by trained personnel, and require active participation from the person being assessed, resulting in reduced access and scalable.

Digital transformation of neuropsychological tests, such as digital trail-making test [5] and digital clock drawing test [6], has been proposed as a way to address these limitations in administration and data recording.

Another approach to address the scalability issues is to design new digital-native neuropsychological tests. These digital tasks also enable assessment of many features that reflect a variety of cognitive and temporal processes that go far beyond a single score, often with the help of signal processing and machine learning approaches. One heavily explored direction is the assessment of cognitive functions and oculomotor abnormalities by examining the visual exploration behaviors during cognitive tests. Both alterations of basic oculomotor function and complex viewing behavior have been observed in people with cognitive impairment (CI) [7].

Over the last two decades, various tasks have been proposed to assess fundamental oculomotor changes in saccades, smooth pursuit, and pupillary responses in people with CI [7]. Both prosaccade (also known as visually guided saccade) and voluntary antisaccade tasks have differentiated CI individuals from healthy controls, where CI individuals showed higher latency and error rates in performing both types of saccade-related tasks [8]. Saccadic intrusions were found to be more frequent in CI individuals [9]. Smooth pursuit deficits have also been identified [10], in the form of lower initial acceleration, decreased velocity, and erroneous anticipatory saccades in the direction of target motion. Studies have also shown that CI individuals have reduced amplitude [11], increased latency [12] of light-induced pupillary changes, and diminished memory-related pupillary responses [13]. CI-related deficits also include disengaging and reorienting spatial attention [14], [15], inhibitory dysfunction due to dorsolateral prefrontal cortex degeneration [16], cognitive decline in attention, and working memory [17].

Other eye-tracking tasks that aim to assess more complex, top-down viewing abilities directly related to cognitive processing have been less explored than tasks that assess fundamental ocular functions. The most widely adopted task of this type is visual search, where CI individuals were found to spend more time and more numerous, dispersed fixations to detect the target objects [18], [19], especially in repeated scenes [13], compared to healthy controls, which may indicate deficits in attention and visual processing. Another task exploits scene exploration, primarily focusing on measuring the viewing behavior in the presence of incongruent scenes. However, the results from this task were mixed, where both lower interest [20] and higher interest [21] in the incongruent scenes were found.

Eye-tracking tasks provide automatic and time-efficient assessments for oculomotor and cognitive functions. However, there remain some challenges. First, most tasks require active participation, which could elicit negative experiences similar to traditional neuropsychological tests due to anxiety and/or perceived poor performance. Second, task administration often requires standalone eye-tracking devices, which increases the cost and reduces accessibility. Lastly, most tasks do not assess the multiple oculomotor and cognitive functions involved in the visual exploration process, hence costing additional time to administer and missing the opportunity to assess visual exploration.

To address the these challenges, in recent work, we developed a 4-min fully-passive mobile-based eye-tracking task to assess visuospatial memory [22], [23], where participants went through free-viewing sessions with camera-based eye-tracking. We have also expanded the analyses to the expressions of emotions during this task and provide an additional assessment of emotional state [24], which is closely connected to cognitive functions. With these features, we screened general cognitive impairment (defined by a score of less than or equal to 24 in MoCA) with good accuracy (with an area under the receiver-operator curve of 0.77) and assessed multiple functions in a single test.

The aim of the current study is to develop a framework that provides an improved automated cognitive impairment screening method and additional quantitative assessments of multiple oculomotor and cognitive functions, without changing the administered VisMET task. More specifically, we aimed to extract largely mutually-independent features from the estimated eye-gaze locations on the corresponding pictures displayed and hypothesized that each feature could provide independent information for the characterization of various cognitive domains. Oculomotor features such as fixation, saccade, and pursuit properties would also be extracted to benchmark the proposed novel features.

The first aspect we were interested in is the spatial distribution of the eye-gaze locations. We hypothesized that differences in spatial distribution could be found in CI individuals, such as more numerous and dispersed distributions, because of their deficits in spatial attention [14], [15].

The second aspect we targeted was the spatiotemporal dynamics of eye-gaze movement. We hypothesized that the declined spatial attention [14], [15] and working memory [13], [17], inhibitory dysfunction [16] in CI may lead to differences in temporal dynamics, such as more or less frequent jumps between regions of interests (ROIs).

The third aspect we aimed to study was whether the intrinsic meaning of the images, i.e., their semantic value, influenced viewing performance. Various semantic categories of images may be processed differently as a result of CI. .For this purpose, we analyzed how different categories of images (e.g., people, animals, objects, and background), influence image viewing patterns, including the quality and preferences of an individual’s semantic organization as well as image contents. Stable individual differences in salience along a set of fundamental semantic dimensions were found in a previous study, which indicates that the semantic preferences may reflect the features of the observer [25]. Hence we hypothesized that CI might alter semantic preferences due to deficits in semantic memory [26].

We evaluated the proposed framework in participants, including healthy controls and patients with MCI or AD, and use the resulting assessments to provide additional overview of the visual exploration deficits manifested in cognitive impairment.

## II. Dataset

### A. Participants

We recruited 348 participants from the Emory Healthy Brain Study (EHBS, n=169) and the Goizueta Alzheimer’s Disease Research Center-affiliated clinics (ADRC, n=179) at Emory University. The ADRC participants consisted of participants with varying levels of cognitive impairment due to AD or related disorders.

Table I shows the demographics of the participants. All procedures followed were in accordance with the ethical standards of the responsible committee on human experimentation.

**TABLE I:** Demographics of the 348 Participants Grouped by Study. ± indicates the standard deviation of the measured variable. The year of education indicates the number of academic years a person completed in a formal program provided by elementary and secondary schools, universities, colleges, or other formal post-secondary institutions. High school completion usually corresponds to 12 years of education, whereas completion of college usually corresponds to 16 years of education. Participants with a total MOCA score greater than 24 were considered cognitively unimpaired, and a MoCA score less than or equal to 24 indicated cognitive impairment.

|  | EHBS | ADRC |
| --- | --- | --- |
| Number of Subjects | 169 | 179 |
| Age (Years) | 69.7 ± 9.3 | 72.3 ± 9.0 |
| Years of Education | 16.6 ± 2.0 | 16.0 ± 2.7 |
| MoCA Score | 25.4 ± 4.0 | 17.8 ± 6.9 |
| Cognitively Unimpaired/ Impaired | 116 / 53 | 33 / 146 |

#### Capacity to provide consent

Special considerations are necessary for adults with Alzheimer’s disease and related disorders that affect cognitive abilities and thus have the potential to impair an individual’s capacity to understand and provide consent. To address this concern, we ensured that the individual(s) signing the assent/consent form, whether the participant or the participant’s representative, had a full understanding of the study. Those providing consent were asked to reiterate what they understood to be the primary goal of this study, the risks, benefits, and requirements of participation. Although many participants with Alzheimer’s disease could provide consent independently, dual consent was obtained from all participants unable to independently provide consent by consenting both the individual with a diagnosis as well as their legally authorized representative prior to enrollment in the study. The consent procedure and this study have been formally approved by the Emory University Institutional Review Board (IRB00078273).

### B. Measurements

All participants received neuropsychological evaluations comprised of many cognitive measures intended to comprehensively measure global cognition as well as specific domains of cognitive function. The MoCA (version 8.1) [2] was used as a standard screening measure to evaluate global cognitive performance in both the EHBS and ADRC cohorts. The MoCA score ranges from one to 30, where only integer scores can be obtained. Participants with a total MOCA score greater than 24 were considered cognitively unimpaired (CU), and a MoCA score less than or equal to 24 was indicative of CI. Neuropsychological tests including Benson complex figure copy (immediate and delayed) [3], forward and backward digit span (DS) [4], F-A-S fluency [27], animal category fluency [27], Trail making tests (part A and B) [28] and Consortium to Establish a Registry for Alzheimer’s Disease (CERAD) word list recall (delayed) [29] were administrated to most participants. Due to administrator error, patient fatigue, or other unforeseen problems, some of the tests could not always be administrated. The number of participants who were administered each test can be found in Table III.

**TABLE II:** Classification performance of cognitive impairment. The term *all* represents performance using all images (including those that were modified), and *og* indicates the use of only the original unmodified images.

| Feature type | AUROC-all | F1-all | AUROC-og | F1-og |
| --- | --- | --- | --- | --- |
| 1. Oculomotor features | 0.64 | 0.60 | 0.58 | 0.54 |
| 2. Spatial distribution (spatial) | 0.68 | 0.63 | 0.62 | 0.57 |
| 3. HMM transition matrix (temporal) | 0.64 | 0.69 | 0.59 | 0.65 |
| 4. All HMM features (spatiotemporal) | 0.69 | 0.70 | 0.64 | 0.65 |
| 5. Semantic viewing time (semantics) | **0.73** | **0.74** | 0.65 | 0.61 |
| 6. Modification viewing time (memory) [23] | 0.73 | 0.72 | NA | NA |
| 7. Combined (all: 2+4+5) | **0.78** | **0.76** | NA | NA |

**TABLE III:**
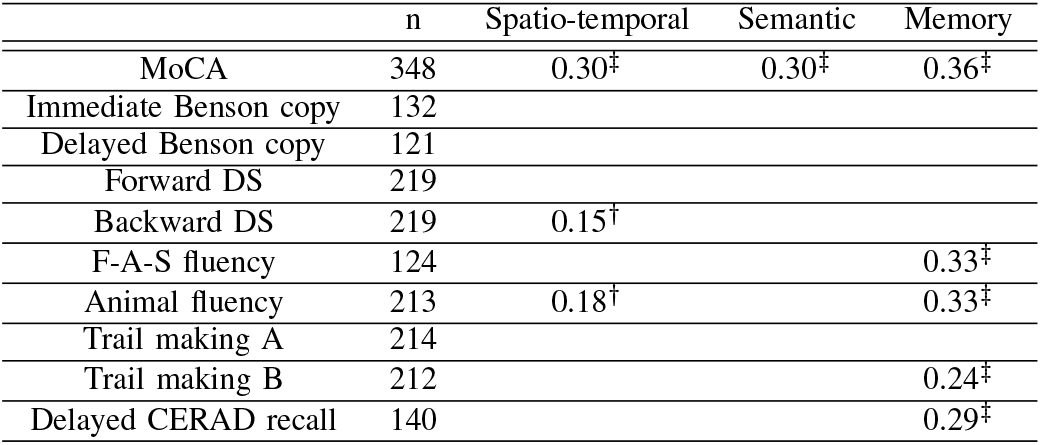
R squared scores of different features on neuropsychological tests. Values in the second column (“n”) show the number participants with the test administered. Values in other columns are the R-squared values between the ground truth scores and the predicted scores in testing folds for each neuropsychological test. A † over the R squared value indicates a significant correlation (*P <* 0.05) between them, where a ‡ indicates that *p <* 0.01. R-squared values were omitted in pairs where no significant correlation was found.

|  | n | Spatio-temporal | Semantic | Memory |
| --- | --- | --- | --- | --- |
| MoCA | 348 | 0.30‡ | 0.30‡ | 0.36‡ |
| Immediate Benson copy | 132 |  |  |  |
| Delayed Benson copy | 121 |  |  |  |
| Forward DS | 219 |  |  |  |
| Backward DS | 219 | 0.15† | | |
| F-A-S fluency | 124 | | | 0.33‡ |
| Animal fluency | 213 | 0.18† | | 0.33‡ |
| Trail making A | 214 |  |  |  |
| Trail making B | 212 | | | 0.24‡ |
| Delayed CERAD recall | 140 | | | 0.29‡ |

### C. Data collection

The Visuospatial Memory Eyetracking Test (VisMET) is a passive viewing test, with participants simply asked to view and enjoy the images displayed on the screen. The participants were not asked to remember the images and did not get scores or feedback during the test. The details of the test were described in [22], [23]. In short, the task first shows ten images of scenes consisting of two to five objects for five seconds, then displays a modified set of images with either one object added or removed from each image. Then the task repeats the process described above by showing another ten different images and ten modified images. The same protocol described in [23] was followed for memory test administration.

A mounted iPad Air 9.7” tablet with maximum screen brightness was used to present the test in portrait orientation. Each iPad was running at least iOS 10. The videos were captured from the tablet’s forward-facing camera at a resolution of 720p and a sampling rate of 30Hz. The clinical testing rooms where the data were captured had natural lighting from windows and overhead fluorescent or LED bulbs. During the calibration procedure, the participants were instructed to move their position to fit the silhouette of a face that appeared on the screen, resulting in an approximate distance of 350 mm between the iPad and the participant’s eyes. The facial video from each participant during the memory test was collected, resulting in a dataset of 348 videos.

### D. Computer-vision-based eye tracking

A deep learning-based eye-tracking method was used to estimate the eye-gaze location of the participant at each frame. After that, the estimated eye-gaze locations were combined into the scanpath (the eye-gaze locations over time during the test) for each participant.

The details of this framework can be found in “Method 4” proposed in [23]. In short, the processing pipeline consists of: 1) a regression tree-based face and eye detection and cropping; 2) a convolutional neural network (CNN) for eye-gaze location estimation trained on MIT’s GazeCapture dataset [30] and on all the videos collected from our previous Emory study [31]; 3) a support vector regression (SVR) layer for eye-gaze estimate calibration trained for each video; and 4) a recalibration of the SVR eye-gaze estimation using a fixation cue between each image. The average test error between the eye-gaze location estimate and the target in the test set was 1.98 cm on a 9.7” (24cm x 16.95cm) display.

## III. Methods

Distinct types of features were extracted from the estimated eye-gaze locations on the corresponding pictures displayed during the test. Then, these features were evaluated in two analyses: 1) the classification of cognitive impairment and 2) the regression on neuropsychological tests. Additional statistical analyses were used to investigate the potential deficits found in CI individuals. The illustration of the processing pipeline can be found in Fig. 1, while the details are described in the following sections.

**Fig. 1:**
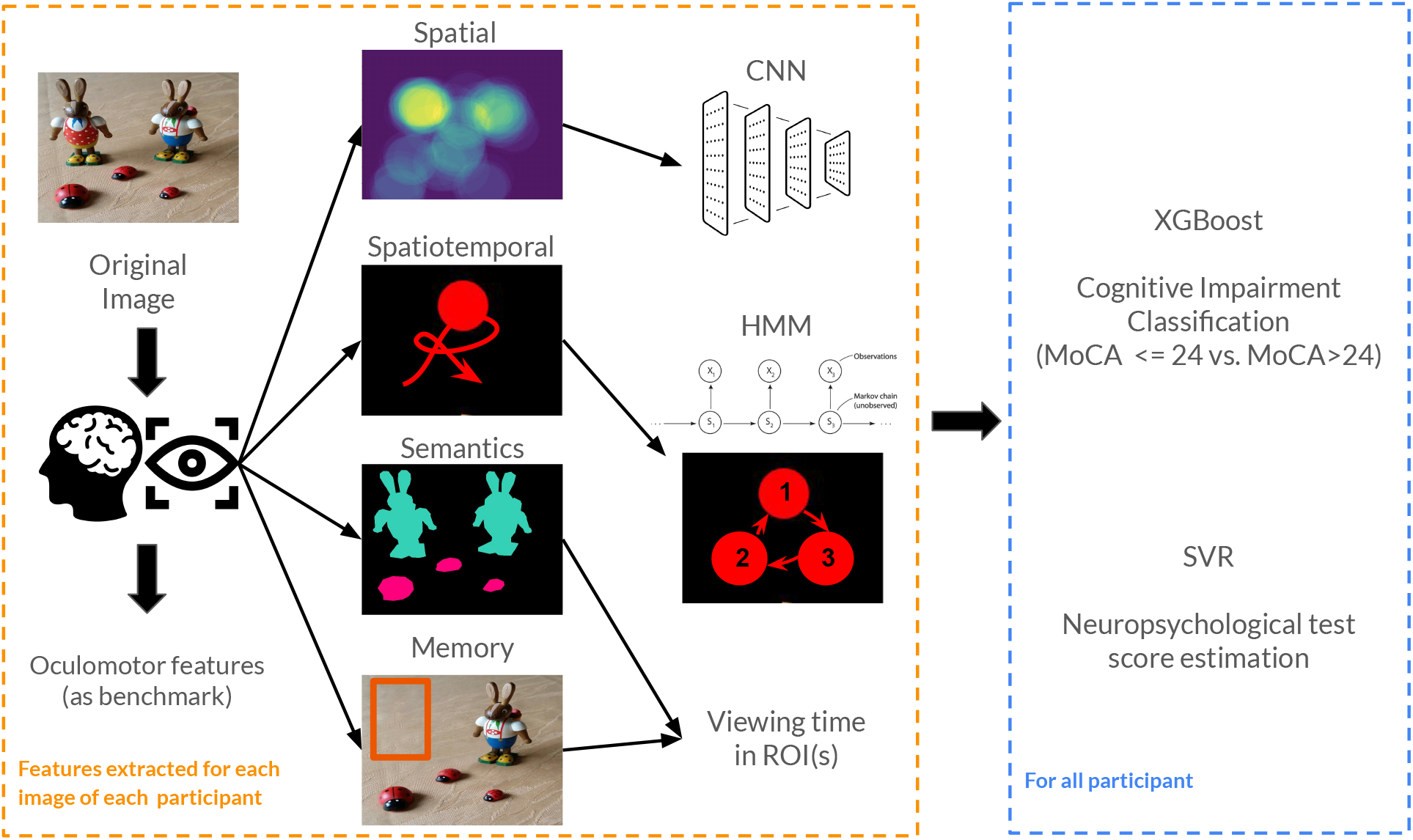
Overview of the processing pipeline. Features with different information were extracted from the scanpath of each image viewed by each participant and then used for cognitive impairment classification and neuropsychological score estimation. Spatial distribution of eye-gaze locations was used to train a convolutional neural network (CNN) for single-image cognitive impairment classification. Then, the single-image predictions of all images were used as features for the final subject classification. Hidden Markov Models (HMM) were trained in an unsupervised manner with the scanpath, and the properties of the HMMs were used for both classification and score estimation. The viewing time of different semantic categories (coded in different colors only for visualization) and the modified region (showed in area within the red bounding box) were used to approximate the semantic preferences and the visual memory capability, which were then used as features for classification and score estimation. Eye movement event were also detected from the estimated gaze and oculomotor features were extracted and used as a benchmark to the novel features proposed in the study. Extreme gradient boosting (XGBoost) [32] was used as the classifier for all features, while support vector regression was used to estimate scores.

### A. Feature generation

#### 1) Spatial distribution of the scanpath

For each fivesecond image viewing session of each participant, a 180×240 heatmap (scaled 25x smaller than the original 900×1200 resolution used in the display) was generated from the estimated scanpath. At each frame (or time point), a filled circle with a center at the estimated eye-gaze location and a radius of the estimation error was added to the heat map. Then the map was normalized so that the sum of all values in the map is one.

#### 2) Spatiotemporal modeling of the scanpath

Since temporal dynamics cannot be modeled directly without the use of spatial information, a hidden Markov model (HMM) was used to model the spatiotemporal dynamics of the scanpath. We hypothesized that there are hidden ROIs that the participant transitioned back and forth to during the free viewing session. Based on the images’ complexity, we set the number of states (ROIs) to be the number that maximized the log-likelihood and the maximum number of states to be three.

For each image viewed by each participant, we learned one HMM. We extracted parameters of the HMM as features, including the prior of the states, transition matrix between different ROIs, ROI center locations, and covariance matrix using the “Scanpath Modeling and Classification with Hidden Markov Models Toolbox” [33]. Unlike the rest, the transition matrix was considered a temporal feature since it represents how participants alternate between ROIs. Additionally, we calculated the multiscale entropy of the inferred states time series of each image viewed by each participant to characterize the complexity of their viewing dynamics, using the “PhysioNet Cardiovascular Signal Toolbox” [34]. Time series of inferred states were scaled into coarse-grain time series by taking the mode of multiple states.

#### 3) Viewing time of different semantic contents

Objects and backgrounds in all 40 images shown during the test were manually annotated with semantic category and boundaries. That is, a semantic category was assigned to each pixel of the images. The semantic category include “background”, “text”, “objects” (such as bicycles, cars, computers), “animal” (such as birds, dogs, rabbits), and “people”. Since the gaze veer away from the screen, we additionally define this scenario as a semantic type of “off-screen”.

The percentage of time a participant spent in a semantic type during a five-second image session was calculated as the average ratio of the overlapping area between the semantic region and the estimated gaze circle (a filled circle with a center at the estimated eye-gaze location and a radius of the estimation error) to the area of the gaze circle at each frame. The average percentages of time the participant spent on different semantic types during the entire test (during all 40 images) were used as the semantic preference features.

#### 4) Viewing time of the modified region

We followed the definition proposed in [23] and defined “modification viewing time” as the percentage of frames (excluding the ones without face/eyes detected) where the estimated gaze is in a fixed expert-defined elliptical region (for each object/picture). More details can be found in our previous studies [23], [24].

#### 5) Oculomotor features

Traditional eye movement events were also extracted from the scanpath of each image viewed by each participant with the “REMoDNaV: robust eye-movement classification for dynamic stimulation toolbox’’ [35]. Various event-related features were generated from those detected events and were evaluated with the rest of the features in the later classification or regression analyses as benchmarks. Those event-related features include (1) the number of different eye movement events: fixation, pursuit, saccade, post saccadic oscillations (PSO); (2) duration of the fixation; (3) the amplitude, velocity, angle, and duration of the saccade.

### B. Classification of cognitive impairment

We evaluated features generated from the above-described processes in a task of the classification of CI vs. CU. Classification performances were measured by the average area under the receiver operating characteristic (AUROC) and the average F1 score (the harmonic mean of the precision and recall) in 20 repeated five-fold cross-validations, where subjects were randomly stratified into five approximately equally sized folds in each repetition.

A two-stage classification approach was used for spatial features (distribution of the scanpath). The spatial distribution maps were first trained at the image level, where distribution maps from participants in the training split were used to train a CNN (EfficientNet [36]) for CI vs. CU classification. On the test fold, the CNN-predicted labels of all images were first combined at the subject level and then used as features for the subject-level classification. For all other types of features, single-stage classification with extreme gradient boosting (XGBoost) [32] was used as the classifier.

To disentangle the potential memory deficits from the rest of the viewing differences, we further evaluated the classification performance only in sessions where the original images were shown (i.e., when the participants explored those images for the first time).

### C. Regression on neuropsychological tests

Different sets of features were evaluated in a task of the regression on scores from different neuropsychological tests, including MoCA, Benson complex figure copy (immediate and delayed), forward and backward DS, F-A-S fluency, animal category fluency, Trail making tests (part A and B) and delayed CERAD. Support vector regression (SVR) was used as the regressor for all the features. Similar to the classification task, 20 repeated five-fold cross-validations were used. The average R-squared value between the neuropsychological test scores and the estimated scores in the 20 testing folds was used as the regression performance metric. Three types of features were evaluated, including spatiotemporal features generated using HMM modeling, semantic features that summarized the viewing time of different semantic contents, and memory features that summarized the viewing time of the modified region.

### D. Statistical Analyses

We used statistical tests to assess the differences in the probability distributions of features between CI individuals and healthy controls and the differences in performance resulting from different features. The Shapiro-Wilk test was used to confirm whether the distributions were normally distributed or not. If not, a two-sided Mann-Whitney rank test was applied between semantic viewing times derived from subjects assessed as CI or CU to determine whether a significant difference exists between these two averages. McNemar’s test was used to evaluate the classification disagreement between pairs of classification settings. Wald Test was used to determine if a significant correlation was found between the estimated values and the actual test scores. Significance was assumed at a level of *p <* 0.05 for all tests.

## IV. Results

### A. Spatially centered gaze in CI individuals

Fig. 2 illustrates the average spatial distribution of eye-gaze locations in CI individuals and healthy controls when viewing the first five original images, along with spatial differences between the two groups. While gaze from both groups primarily focused on the center and salient objects of the images when viewing for the first time, people with cognitive impairment spent more time in the center area of the image, characterized by the dark circular regions found in the figures. Significantly higher (Mann-Whitney test, *p <* 0.01) average pixel values in the central area (a circle with a radius of the eye-tracking estimation error) were found in CI individuals.

**Fig. 2:**
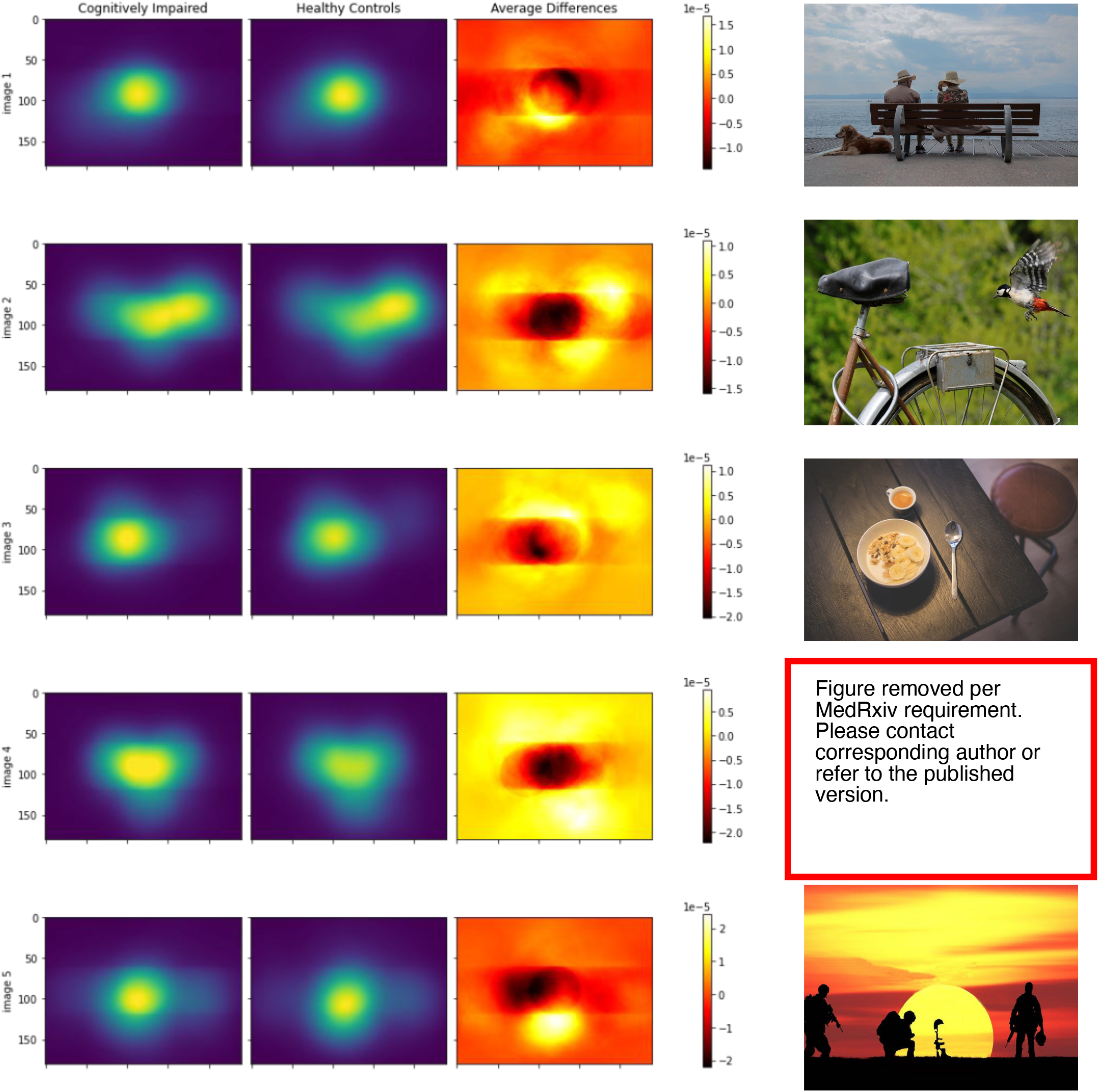
Average spatial distribution of eye-gaze locations in CI individuals, healthy controls, and their differences in the first five original images. The first column of figures shows the average eye-gaze location distribution in participants with cognitive impairment, while the second column shows that in healthy participants. The first two heatmaps on each row were plotted with the same range of color, with the value ranging from zero to the maximum in all participants. The third column showed the average differences between the first two columns (healthy controls - CI individuals). Figures in the first two columns were plotted with a color map different from the one used to plot the heatmap in the third column, but brighter colors in both color maps indicate large values, i.e., more viewing time.

### B. HMM statistics

Statistics of the learned HMMs in the 20 original images revealed significant differences between CI individuals and healthy controls when they explored those images for the first time. On average, people with cognitive impairment tend to view a greater number of ROIs (Mann-Whitney test, *p <* 0.01) compared to healthy controls. In 69% of images, CI individuals viewed three ROIs, 22% viewed two ROIs, and 9% viewed one ROI. In comparison, in 66% of images, healthy controls viewed three ROIs or more, 23% viewed two ROIs, and 11% viewed one ROI.

The Fig. 3 (left panel) visualizes the average transitioning probability modeled by the HMMs, where people with cognitive impairment showed a significantly lower probability (Mann-Whitney test, *p <* 0.01) switching between different ROIs.

**Fig. 3:**
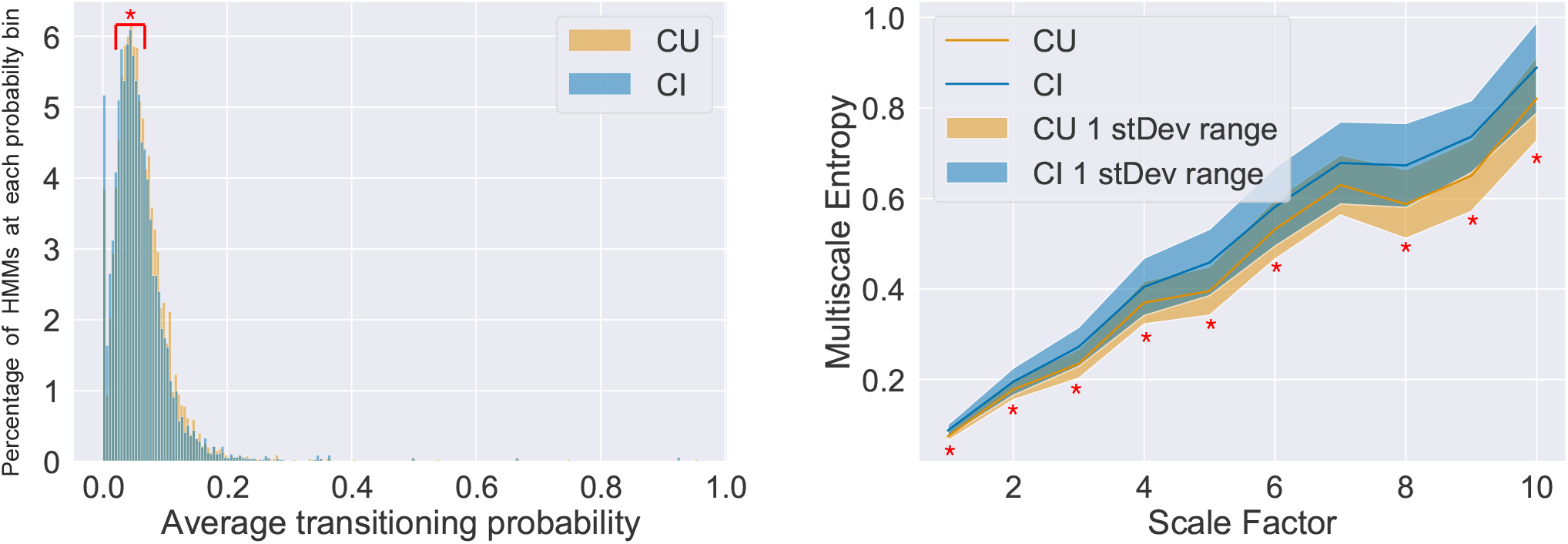
Inferred hidden Markov model characteristics when viewing original images. Red stars (“*”) indicate statistically significant (Mann-Whitney test, *p <* 0.05) differences between healthy controls (cognitively unimpaired or CU, in yellow) and cognitively impaired participants (CI, in blue). **(Left)** Histogram of average transitioning probability in all participants, showing the distributions of the average probability of going from one state to another different state learned by hidden Markov models from all original images. **(Right)** Multiscale entropy of inferred states in all participants. Sample entropy of hidden states time series inferred by the hidden Markov model was calculated at the scale of one (i.e., without scaling) to ten. The average entropy and the one standard deviation range are shown.

While CI individuals switched between ROIs less often, their alternating pattern showed higher complexity when they did. The right panel in Fig. 3 shows the average and one standard deviation of sample entropy measured at different scales. In almost all scales (except for seven), CI individuals showed a significantly higher level of entropy (Mann-Whitney test, *p <* 0.05).

### C. Semantic class preference alternation during first and second exploration

Fig. 4 shows the distribution of viewing time of semantic categories with significant viewing time differences between CI individuals and healthy controls in modified and original images.

**Fig. 4:**
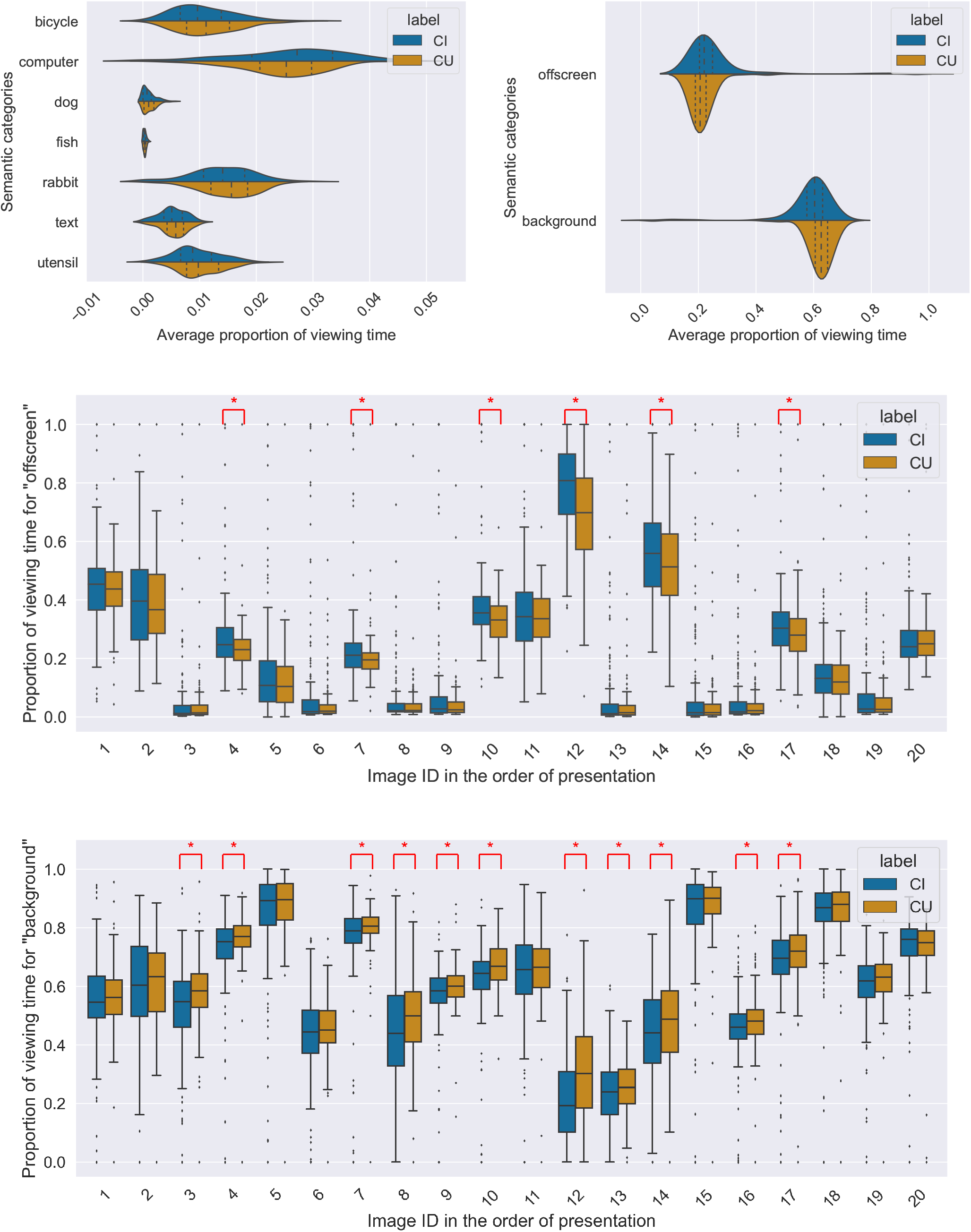
Semantic categories with significant viewing time differences between CI individuals and healthy controls in modified and original images. **(Top Left)** Average proportion of viewing time spent in categories with significant differences between CI individuals and healthy controls in original images. **(Top right)** Average proportion of viewing time spent in “off-screen” and in “background” in modified images. **(Middle)** Percentage of viewing time spent in “off-screen” and **(Bottom)** in “background” in each of the modified images. A red star indicates significant differences between CI individuals and healthy controls in that image.

Significant differences were found in the exploration of the seven semantic categories of images during the first presentation (the original images), shown in the Fig. 4 (top left panel). CI individuals consistently spent less time viewing animals, and less time viewing text, and had a mixed effect for time spent on objects, indicating semantic preferences might be associated with cognitive impairment, potentially due to semantic memory-related deficits.

During the second presentation with the modified images, shown in the top right sub-figure of Fig. 4, there were significant differences in the exploration of many categories of images. However, it is worth noting that on average, CI individuals spent significantly more time off-screen and significantly less time in the background, suggesting decreased attention to non-salient image components. The middle and bottom panels of Fig. 4 further show that the pattern we identified in different categories still holds in almost all the individual images.

### D. Performance of cognitive impairment classification

Table. II shows the classification performance of cognitive impairment using different types of features. All features proposed in this work outperformed the benchmark oculomotor features in CI vs. CU classification statistically significantly (*P <* 0.01), which suggests additional information in those features beyond eye movement characteristics. This result aligns with the significant differences we found between groups in the above sections. Semantic viewing time features resulted in the best performance of an AUROC of 0.73 with a single type of feature while combining it with spatiotemporal and memory features improved the performance statistically significantly (*P <* 0.01) to an AUROC of 0.78.

### E. Performance of neuropsychological test score estimation

While good performance in cognitive impairment classification and significant correlations were found with MoCA, suggesting all features are cognitively relevant, the proposed features cannot fully account for all cognitive functions measured in the neuropsychological battery. That said, spatiotemporal features and memory features were significantly correlated with specific measures, as shown in Table. III.

The ability to use spatiotemporal features to estimate backward digit span scores suggests that those features are tapping into aspects of working memory and complex attention by measuring where the participants were looking and how they alternate between the ROIs. Spatiotemporal features were also associated with animal fluency, which is a semantic verbal generation task that relies both on semantic knowledge and executive functioning performance. This association may suggest that image switching is associated with more semantically relevant aspects of executive function, since it is not associated with other more concrete executive functioning tasks (such as Trail Making Test B, a measure of set shifting).

Memory features correlated with multiple measures, which span executive function, verbal generation, and delayed memory recall. Specifically, memory scores were significantly associated with F-A-S fluency and animal fluency, both of which rely heavily on frontal executive circuits for task performance. In line with this, Trails Making Test B performance was associated with this feature, again suggesting executive function is related to these features. These features were also, as expected, associated with Delayed Recall on the CERAD, a measure of memory function. Although these features are meant to capture memory function, these findings suggest that both memory and executive function are heavily related to the Memory features. This may be because we are including both added and removed images in task performance. As image components are removed, the ability to recognize that something is missing from a previously studied image should be heavily reliant on memory function; however, as image components are added, it may not be the case that attention to these image components relies on memory performance and instead may associate more highly with executive functioning (since recognition of differences is not required for engaging with the critical region).

## V. Discussion and conclusion

We utilized a combination of visual exploration features from participants passively viewing a series of original and modified images to capture cognitive impairment. Using the same passive VisMET viewing task [23] that we developed to tap into memory function, the current methods captures additional features that tap into other cognitive processes. The revised analytical method resulted in improved performance in CI screening and neuropsychological score estimation, as shown in Table. II and Table. III. We also identified differences in the spatial and spatiotemporal processing of participants with cognitive changes, as well as in the viewing pattern of objects based on their semantic types. These features were identified when participants viewed the original images for the first time, which showcased how they explored novel scenes proactively.

People with cognitive impairment are known to have deficits in spatial attention [14], [15]. The results of our study further confirmed and depicted a more fine-grained landscape of the attention deficit in cognitive impairment. Spatially centered attention was observed in CI individuals (shown in Fig. 2). Additionally, CI individuals had more ROIs than controls yet did not spend more time wandering in the background or off the screen. Those findings indicate that CI individuals might have a spatially misdirected attention rather than just a generally reduced attention. Similar to the difficulty in disengaging and reorienting spatial attention [14], [15] and inhibitory dysfunction [16] shown during active tasks found in previous literature, CI individuals in our study had a slightly lower average probability of switching ROIs (shown in Fig. 3) when they were passively exploring without instruction. They also showed more chaotic ROI transitions (characterized by higher entropy, shown in Fig. 3). Unlike active tasks, it is safe to say that those behaviors observed in our passive test were not a result of the difficulty in following instructions but a fundamental deficit in spatial attention.

The altered preferences for various semantic categories of objects shown in Fig .4 suggest higher-level changes in conceptualization and understanding of objects or scenes in individuals with CI. When they viewed the similar but modified scenes for the second time, they showed a pattern of spending more time in the background and wandering their gaze off the screen. The increased time spent off the screen might result decreased persistence in attention, while the decreased time spent in the background may reflect decreased attentional resources and in general is consistent with the aforementioned lack of persistence in attention wherein participants spent more time engaged in “off screen” viewing across the task. Surprisingly, this difference was not seen when individuals viewed the original set of images, and suggests that possibly individuals with CI had fatigued and/or generally lost attention during the latter two minutes of the task.

Although we found that scene memorability is a property of pre-existing semantic associative in long-term memory in healthy controls [37], we did not observe semantic-specific memory deficit at the group level in this study. This finding may indicate that short-term semantic memory change might also be individualized, in contrast to the observed group-level semantic preference differences manifested when viewing the original images in this study. These group-level differences in semantic valence might suggest that semantic knowledge related systems weaken in people with CI, hence they may be less efficient in grouping or less coherent in how they approach images.

We further confirmed that the spatial, spatiotemporal, semantic, and memory features could be used separately or in combination, to classify cognitive impairment with improved performance. By carefully evaluating each aspect of the viewing behavior, we achieved far better results than using only the benchmark oculomotor features, as well as our previous methods using only the memory features [23] and the facial expression features [24].

While an AUROC of 0.78 is promising for a four-minute passive test, there are several additional factors that could potentially be optimized to improve performance of the task.. For example, the selection of images and the corresponding objects included was not optimized on the ability to differentiate CI individuals from healthy controls. We have found that the CI vs. CU group differences were drastically different in different images, which could be leveraged for image optimization. Some images showed clear separation in multiscale entropy level between two groups, while others had two groups almost overlapping. Group differences in semantic viewing time also varied in different images, like the viewing time spent in the background and offscreen shown in Fig. 4. Clearly, we cannot simply optimize based on how well each image performs in this dataset and implicitly overfit the model to this dataset. However, extrapolation can be made, on top of our interpretation of the mechanism, to help decide which images, objects should be included in the tests and how they should be presented to maximize the power of the test.

Based on the significant group differences and the ability to classify cognitive impairment, spatiotemporal, semantic, and memory features clearly measured certain aspects of general cognitive ability. Unfortunately, their relations with existing neuropsychological tests are less clear. For instance, spatiotemporal features correlated with semantic and verbal functions, yet surprisingly semantic features did not correlate with these functions as shown in Table. III. Similarly, none of those three feature types could effectively estimate the scores of Benson copy, which measures visuoconstructional and visual memory functions, which is expected to correlate with spatiotemporal and memory features intuitively. Those results suggest that further investigation is needed to validate which cognitive functions are measured by either the visual exploration features proposed in this work or the traditional neuropsychological tests. It could be the case that those proposed features are measuring functions that the traditional tests were not measuring, yet we currently do not have the data to prove it. It is also worth noting that we have significantly fewer subjects with specific neuropsychological tests administered compared to MoCA, which is one of the reasons why the estimation of the scores was more difficult than estimating MoCA: it is more difficult to learn an effective estimator with fewer data points.

## Data Availability

Raw data cannot be shared publicly since it contains personal identifiable information (video recordings of participants' faces). Although participants agreed to be recorded for purposes of the work, there are restrictions in place for the release of PII. Additionally, due to the sensitive nature of the data that we collect, Certificates of Confidentiality are in place to prohibit disclosure of identifying information

## VI. Acknowledgements

This research was supported by funding from the James M. Cox Foundation, the Goizueta Foundation and the Goizueta Alzheimer Disease Research Center at Emory University (P50 AG025688), and the Emory Healthy Brain Study (R01 AG070937).

